# Perceptions, views and practices regarding antibiotic prescribing and stewardship among hospital physicians in Jakarta, Indonesia

**DOI:** 10.1101/2021.09.05.21263144

**Authors:** Ralalicia Limato, Erni J. Nelwan, Manzilina Mudia, Monik Alamanda, Elfrida R. Manurung, Ifael Y. Mauleti, Maria Mayasari, Iman Firmansyah, Roswin Djafar, Vu Thi Lan Huong, H. Rogier van Doorn, Alex Broom, Raph L. Hamers

## Abstract

**Objectives:** Antibiotic overuse is one of the main drivers of antimicrobial resistance (AMR), especially in low and middle-income countries. This study aimed to gain an understanding of perceptions, views, and practices regarding AMR, antibiotic prescribing, and stewardship (AMS) among hospital physicians in Jakarta, Indonesia.

**Design:** cross-sectional, self-administered questionnaire-based survey, with descriptive statistics, exploratory factor analysis (EFA) to identify distinct underlying constructs in the dataset, and multivariable linear regression of factor scores to analyse physician subgroups.

**Setting:** Six public and private general hospitals in Jakarta in 2019.

**Participants:** 1007 of 1896 (53.1% response rate) antibiotic prescribing physicians.

**Results:** EFA identified six latent factors (overall Crohnbach’s α=0.85): awareness of AMS activities; awareness of AMS purpose; views regarding rational antibiotic prescribing; confidence in antibiotic prescribing decisions; perception of AMR as a significant problem; and immediate actions to contain AMR. Physicians acknowledged the significance of AMR and contributing factors, rational antibiotic prescribing, and purpose and usefulness of AMS. However, this conflicted with reported suboptimal local hospital practices, such as room cleaning, hand hygiene and staff education, and views regarding antibiotic decision-making. These included insufficiently applying AMS principles and utilising microbiology, lack of confidence in prescribing decisions, and defensive prescribing due to pervasive diagnostic uncertainty, fear of patient deterioration or because patients insisted. Physicians’ factor scores differed across hospitals, departments, work experience and medical hierarchy.

**Conclusions:** AMS implementation in Indonesian hospitals is challenged by institutional, contextual and diagnostic vulnerabilities, resulting in externalising AMR instead of recognising it as a local problem. Appropriate recognition of the contextual determinants of antibiotic prescribing decision-making will be critical to change physicians’ attitudes and develop context-specific AMS interventions.

**Strengths and limitations of this study:**

- The self-developed questionnaire in this study identified a relevant set of attributes through a factor analysis optimization process, with adequate content, face and construct validity and internal reliability. This study adds important value in the absence of adequately validated instruments regarding antimicrobial resistance and stewardship, with particular applicability for LMIC.
- This study had a large, varied respondent sample and high response rate among physicians at six public and private hospitals in Jakarta, Indonesia, and identified differences between physicians across hospitals, departments, work experience and medical hierarchy, which can guide priority-setting and tailoring of stewardship interventions.
- However, non-participation and the convenient hospital sample could have introduced selection bias, and the data are not necessarily representative for Jakarta or Indonesia.
- Factor analysis is based on using a “heuristic”, which leaves room to more than one interpretation of the same data and cannot identify causality.

## Introduction

The global rise in drug-resistant infections is one of the leading threats to public health globally, with increasing rates of morbidity, mortality and escalating healthcare costs^1^. Misuse and overuse of antimicrobial drugs in human health care is one of the main drivers^2,3^ and also represents a key solution, i.e. judicious use of remaining antibiotics. Globally, use of antibiotics remains largely unrestrained and poorly governed, with large, unregulated healthcare systems representing an increasingly challenging area for achieving the goal of optimization. Substantial variations in contributing factors to inappropriate antibiotic prescribing exist across contexts, e.g. diagnostic uncertainty, pressure from pharmaceutical industry or patients^4,5^, with the structure and funding of health systems inflecting enactment of optimization strategies, including antimicrobial stewardship (AMS)^6^.

AMS programs aim to control antimicrobial use, and have been associated with reducing hospital-acquired infections, unnecessary healthcare costs, and potentially drug-resistant infections^7–9^. However, AMS programmes in turn may jar with local constraints and practices and have been shown to have limited traction when attempts to implement occur without adequate understanding of context^10^.

The global push to enact effective AMS requires detailed, context-specific data on physicians, given their central role in the complex process of antibiotic prescribing in hospitals, which can inform on how AMR is conceived, how current prescribing is rationalised, and how broad AMS principles may be experienced across contexts and nations^11,12^. Few studies to date have been conducted on this topic in low and middle-income countries (LMIC), with insufficient evaluation of the psychometric properties of their measurement instruments to examine their suitability to the specific context^4,5^. Indonesia, a diverse middle-income country in Southeast Asia with the world’s fourth largest population (275 million), is particularly vulnerable for AMR, driven by dense urban populations combined with rising antibiotic consumption^13^, a decentralised health system^14^, and weakly enforced antibiotic policies^15^, hence promoting inappropriate prescribing and over-the-counter access without a prescription. Despite progress in government policies, AMS is generally in an early stage of implementation^15,16^.

To identify context-specific opportunities for AMS interventions, we conducted a questionnaire-based survey among antibiotic-prescribing physicians in hospitals in Jakarta, Indonesia, to evaluate their perceptions of AMR, accounts of antibiotic prescribing, and views on AMS. We performed exploratory factor analysis (EFA) to evaluate the construct validity and psychometric properties of the questionnaire, identify distinct underlying constructs in the data, and explore differences between physician subgroups.

## Methods

### Study design and setting

We conducted a cross-sectional survey between March and August 2019 among all antimicrobial prescribing physicians at six public and private general hospitals in Jakarta, Indonesia, as part of a mixed-method study to identify targets for quality improvement in antibiotic prescribing practices (EXPLAIN study^17^). The hospitals included two tertiary-care government hospitals and four secondary hospitals, three of which were private hospitals. At the time of the survey, all six hospitals had an AMS programme, albeit at an early stage of implementation.

All qualified physicians prescribing antibiotics on a regular basis working across all clinical departments were eligible to participate, including interns/internship doctors *(magang/dokter internsip)*, general practitioners (*dokter umum*) (GPs), residents (*residen*), specialist/consultant physicians (*dokter spesialis/konsultan*), and others.

### Ethical considerations

The research ethics committee of the Faculty of Medicine University of Indonesia (1364/UN2.F1/ETIK/2018) and the Oxford Tropical Research Ethics Committee (559-18) approved the study, with additional permission from hospital management. As the survey was anonymous, participant consent was inferred when the doctor completed and returned the questionnaire, as explained in the survey introduction.

### Patient and public involvement statement

Patients or the public were not involved in the design, conduct, or reporting of the research.

### Survey questionnaire

We developed a two-page anonymous, self-administered, paper-based questionnaire, which was easy to complete and based on a conceptual framework that included attributes related to prescribers’ perceptions, views and practices. Behavioural theories, good practice recommendations for questionnaire design and existing questionnaires in the literature were reviewed and discussed with several experts. The Clinician Pre/Post Perception Survey of the Greater New York Hospital Association United Hospital Fund^18^ constituted the initial set of items, supplemented with relevant items from other existing questionnaires^19–25^. From a preliminary pool, we selected 69 items, of which 8 items were worded in the negative to address the acquiescence effect. The instrument was translated from English to Indonesian, and back-translated by an independent translator. The questionnaire was pre-tested by a convenience panel of 18 physicians (2 GPs, 15 residents, and 1 consultant), and adjustments were made in accordance with their feedback, reducing the number of items to 40. The final version took about 10 minutes to complete.

The final questionnaire included an explanation of study purpose and completion instructions; 40 short statements (items) to which participants were asked to indicate the extent to which each reflected their own opinion on a 5-point Likert scale, divided into 3 sections: scope of the AMR problem and key contributors; antibiotic prescribing practices; AMS; and respondent socio-demographics (**Appendix**).

### Respondent recruitment

The hospital management provided the total number of prescribing physicians for each department. The questionnaires were delivered to the head of each unit who then distributed the survey to all eligible staff. The study coordinator kept a record of numbers of physicians approached and participated. Upon survey completion, respondents could enter a raffle to win one of three gift cards in each hospital (US$14 each); there were no other incentives for participation.

### Ethical approval

The research ethics committee of the Faculty of Medicine University of Indonesia (1364/UN2.F1/ETIK/2018) and the Oxford Tropical Research Ethics Committee (559-18) approved the study, with additional permission from hospital management. As the survey was anonymous, consent was inferred when the participant completed and returned the questionnaire.

### Statistical analysis

The percentage of respondents selecting each answer choice was calculated using the total number of responses as the denominator. For an EFA, a common lower bound for sample size is 10 cases per variable, suggesting a minimum sample size of 400; to allow for meaningful subgroup comparisons and minimize selection bias, we targeted a >50% response rate and a sample size of >1000 across the six hospitals. We performed EFA to identify underlying distinct constructs, using factor, pcf command in Stata with orthogonal (varimax) rotation. For this analysis, the eight items worded in the negative were reverse-coded, and missing data for categorical variables were treated as a separate category. The Kaiser-Meyer-Olkin (KMO) was calculated to ensure EFA requirements were met. Each item was assigned to a certain factor based on the highest absolute factor loading of the rotated solution, and we then assigned an umbrella term to each factor. After the optimal factor solution had been achieved, we calculated factor scores using the regression scoring method, and Cronbach’s α coefficient to test internal reliability. Using the factor scores (dependent variable), multivariable mixed-effects linear regression was used to analyze physician subgroups (independent variables i.e., hospital sector and care level, grouped departments, work experience and medical hierarchy), while adjusting for possible clustering within hospitals as well as relevant confounders. P-values<0.05 were considered statistically significant. All analyses were done with Stata/IC Version 16.1 (StataCorp, College Station, TX, US).

## Results

### Respondent characteristics

All 1896 antibiotic prescribing physicians at the six hospitals were approached, and 1007 (53.1%) participated in the survey. **Table 1** summarizes the participants’ key characteristics. **Table S1** summarizes the response rates.

**Table 1.** Characteristics of respondents

| Total | 1007 |
| --- | --- |
| Sex <sup>a</sup> |  |
| Female | 477 (47.4) |
| Male | 524 (52.0) |
| Professional hierarchy |  |
| Intern/Internship doctor | 10 (1.0) |
| General practitioner | 113 (11.2) |
| Resident | 500 (49.7) |
| Specialist/consultant | 358 (35.6) |
| Other | 18 (1.8) |
| Professional experience (years) <sup>b</sup> |  |
| <1 | 194 (19.3) |
| 1-5 | 459 (45.6) |
| 6-10 | 136 (13.5) |
| 11-15 | 74 (7.4) |
| 16-20 | 52 (5.2) |
| >20 | 81 (8.0) |
| Grouped departments <sup>c,f</sup> |  |
| Surgery (including subspecialties) | 371 (36.8) |
| Medicine (including subspecialties) | 232 (23.0) |
| Acute specialties | 156 (15.5) |
| Other departments | 244 (24.2) |
| Information sources used to guide prescribing <sup>d</sup> |  |
| Guidelines |  |
| International | 619 (61.5) |
| National | 628 (62.4) |
| Hospital | 656 (65.1) |
| Department/Division | 405 (40.2) |
| Consultation with senior colleague(s) | 472 (46.9) |
| Consultation with microbiologist/infectious disease physician | 523 (51.9) |
| Textbooks | 410 (40.7) |
| Medical journals | 389 (38.6) |
| Pharmaceutical company representative | 34 (3.4) |
| Internet | 115 (11.4) |
| Other | 13 (1.3) |
| *Number of AMR/AMS trainings attended in the past year <sup>e</sup>* |  |
| 0 | 396 (39.3) |
| 1 | 342 (34.0) |
| ≥2 | 168 (16.7) |
| Median (range) | 1 (0, 10) |

Abbreviations: AMR, antimicrobial resistance; AMS, antibiotic stewardship;
Data are reported as n (%) unless indicated otherwise.
Data missing for: <sup>a</sup> 6 (0.60%); <sup>b</sup> 11 (1.1%); <sup>c</sup> 4 (0.40%); <sup>d</sup> 2 (0.20%); <sup>e</sup> 101 (10.0%).
<sup>f</sup> Surgery and surgical subspecialties includes obstetrics/gynaecology (146), surgery (122), orthopaedics (57), ENT (32), urology (14); medicine and medical subspecialties includes medicine (128), neurology (63), pulmonology (15), dermatology (14), cardiology (12); acute specialties includes anaesthesiology (72), emergency (57), ICU (27); other departments includes paediatrics (54), ophthalmology (39), multiple units (33), rehabilitation (32), psychiatry (30), dentist (27), other (29) and unspecified (4).

### Exploratory factor analysis

The KMO was 0.8773 overall and >0.5 for all items, suggesting the data were suitable for EFA. Analysis of the scree plot (**Figure S1**) indicated a case for four factors, whereas the parallel analysis (**Figure S2**) indicated a case for seven factors. The four-factor solution yielded strong factors but explained only 39.9% of the variance and lacked a theoretical basis for one factor. The seven-factor solution contained one factor with only three items that was difficult to interpret; two of these items (Q9 and Q10) did not load well with any factor in various alternative factor solutions and were removed. Therefore, a six-factor model with a clear theoretical basis based on 38 items was deemed most suitable, explaining 47.4% of the variance, with KMO 0.8802 overall and >0.5 for each item (**Table 2**). The six latent factors are (**Table 3**, see **Table S2** for details): 1) Awareness of AMS activities; 2) Awareness of AMS purpose; 3) Views regarding rational antibiotic prescribing; 4) Confidence in antibiotic prescribing decisions; 5) Perception of AMR as a significant problem; and 6) Immediate actions to contain AMR. Internal reliability was excellent for the overall 38-item scale (α=0.85) and factor 1 (α=0.8734) and 2 (α=0.8334), good for factor 3, 4 and 5 (α=0.70 each), and acceptable for factor 6 (α=0.57).

**Table 2.** Summary of the exploratory factor analysis of the six-factor solution (n=973)

| Item # | Original item | Rotated factor loadings |  |  |  |  |  | Uniqueness |
| --- | --- | --- | --- | --- | --- | --- | --- | --- |
|  |  | Factor 1 | Factor 2 | Factor 3 | Factor 4 | Factor 5 | Factor 6 |  |
| Q01 | Antimicrobial resistance is a significant problem in this hospital | 0.2159 | 0.04100 | 0.1389 | 0.0057 | 0.5701 | 0.0416 | 0.6056 |
| Q02 | Antimicrobial resistance is a significant problem in Indonesia | 0.2433 | 0.10730 | 0.3295 | 0.0588 | 0.5742 | 0.0385 | 0.4861 |
| Q03 | A cause of antimicrobial resistance is using too many antimicrobial drugs | 0.1641 | 0.20060 | 0.3993 | 0.0695 | 0.5361 | -0.0462 | 0.4790 |
| Q04 | Lack of hand disinfection by healthcare workers causes spread of antimicrobial resistance | 0.0640 | 0.09020 | -0.2533 | 0.0152 | 0.5725 | 0.1179 | 0.5817 |
| Q05 | Use of broad-spectrum antibiotics can increase antimicrobial resistance when narrower-spectrum antibiotics are available that are equally effective | 0.2150 | 0.05360 | 0.1482 | 0.0005 | 0.5480 | 0.1093 | 0.6167 |
| Q06 | Antibiotic resistance is also a problem outside of the hospital, in communities | 0.2088 | 0.03180 | 0.2463 | 0.0632 | 0.4766 | 0.0432 | 0.6617 |
| Q07 | In this hospital, patient rooms are cleaned according to hospital cleaning protocol once a patient with a multidrug-resistant organism (MDRO) has been discharged | 0.1038 | 0.07530 | 0.0868 | 0.0210 | 0.0378 | 0.6058 | 0.6071 |
| Q08 | Adherence to hand-hygiene protocols is excellent at this hospital | -0.0098 | 0.03640 | 0.0606 | 0.1099 | -0.0830 | 0.6368 | 0.5705 |
| Q11 | Antibiotics are overused in Indonesia | 0.1924 | 0.18310 | 0.5138 | -0.0809 | 0.2489 | -0.2318 | 0.5432 |
| Q12 | Antibiotics are overused in this hospital | 0.0548 | 0.04170 | 0.2094 | -0.4550 | 0.1635 | -0.2381 | 0.6610 |
| Q13 | Microbiology laboratory results are efficiently communicated to the treating physician | 0.1689 | 0.11740 | -0.0915 | 0.0532 | -0.0963 | 0.5116 | 0.6754 |
| Q14 | I regularly refer to/consider the antibiotic susceptibility patterns at this hospital/institution (i.e. the institutional antibiogram) when empirically prescribing antibiotics | 0.0288 | 0.10700 | -0.0292 | -0.0237 | 0.2056 | 0.6115 | 0.5701 |
| Q15 | If medically appropriate, intravenous antibiotics should be stepped down to an oral alternative after three days | -0.0873 | 0.20850 | 0.1141 | -0.0850 | 0.2311 | 0.3617 | 0.7444 |
| Q16 | Restrictions on antibiotics impair my ability to provide good patient care | 0.0460 | 0.18390 | 0.0527 | 0.4031 | 0.0843 | -0.0868 | 0.7842 |
| Q17 | More judicious use of antibiotics would decrease antimicrobial resistance | 0.3010 | 0.07470 | 0.7362 | 0.0820 | 0.0892 | 0.0648 | 0.3429 |
| Q18 | Following evidence-based antibiotic guidelines will help optimize treatment outcomes | 0.2274 | 0.19340 | 0.6565 | 0.1206 | 0.1256 | 0.1851 | 0.4153 |
|  |  | Factor 1 | Factor 2 | Factor 3 | Factor 4 | Factor 5 | Factor 6 |  |
| Q19 | In general, rational antibiotic prescribing for my patients is high on my list of priorities | 0.1845 | 0.18040 | 0.5246 | 0.1574 | 0.1184 | 0.3228 | 0.5151 |
| Q20 | Developing hospital antibiotic guidelines is more useful than applying international guidelines | 0.1803 | 0.00750 | 0.3499 | -0.1335 | 0.2631 | 0.0672 | 0.7534 |
| Q21 | I am often unsure if a patient needs an antibiotic or not | 0.0489 | 0.02600 | 0.0497 | 0.5640 | -0.3364 | 0.1021 | 0.5527 |
| Q22 | I am often unsure which antibiotic to prescribe | -0.0084 | 0.04570 | 0.1256 | 0.5670 | -0.1938 | 0.0781 | 0.6170 |
| Q23 | I will stop antibiotics that others have prescribed in the absence of an appropriate indication | 0.0016 | -0.06200 | 0.1517 | -0.1137 | 0.2090 | 0.3892 | 0.7650 |
| Q24 | Patients with high fever ($\geq 39^{\circ}\text{C}$) must be treated with antibiotics | 0.0077 | 0.13310 | 0.1695 | 0.4794 | 0.2095 | -0.2351 | 0.6245 |
| Q25 | If I am uncertain about the diagnosis of infection, but think it is possible, I feel safer prescribing an antibiotic | 0.0217 | -0.04150 | -0.0877 | 0.6741 | 0.1927 | -0.0086 | 0.4985 |
| Q26 | Fear of patient deterioration or complications leads me to prescribe antibiotics more freely | 0.0113 | 0.00390 | -0.1459 | 0.7092 | 0.0883 | 0.0246 | 0.4672 |
| Q27 | I frequently prescribe antibiotics because patients or their relatives insist on it | 0.1069 | 0.04740 | 0.2869 | 0.6318 | 0.0518 | -0.0100 | 0.5021 |
| Q28 | I am aware that my hospital has an antimicrobial stewardship program (ASP) | 0.2434 | 0.62240 | 0.2918 | 0.0866 | 0.0418 | 0.0110 | 0.4588 |
| Q29 | I understand what the purpose of ASP is | 0.2217 | 0.69570 | 0.2635 | 0.1188 | 0.0092 | 0.0199 | 0.3828 |
| Q30 | ASP improve patient care | 0.2364 | 0.77440 | 0.0873 | 0.0932 | 0.0852 | 0.1241 | 0.3055 |
| Q31 | ASP reduces the problem of antimicrobial resistance | 0.2775 | 0.75320 | 0.0949 | 0.0172 | 0.0906 | 0.1753 | 0.3074 |
| Q32 | ASP reduces this hospital's infection rates | 0.2045 | 0.66700 | -0.1122 | 0.0569 | 0.1283 | 0.1799 | 0.4486 |
| Q33 | Additional staff education on antimicrobial prescribing is needed | 0.5202 | 0.28160 | 0.0917 | -0.0079 | 0.2203 | -0.0284 | 0.5923 |
| Q34 | Regular audit and feedback encourage me to prescribe antibiotics prudently | 0.6151 | 0.35810 | 0.0075 | 0.0064 | 0.1402 | 0.1117 | 0.4612 |
| Q35 | Rapid and accurate diagnostic tests are useful for diagnosis of infectious diseases and guidance on antibiotic therapy | 0.6714 | 0.25760 | 0.1522 | -0.0362 | 0.0386 | 0.0379 | 0.4555 |
| Q36 | To reduce antibiotic overuse in hospitals, implementation of antibiotic restriction (e.g. antibiotic tiers) is a useful measure | 0.6428 | 0.26700 | 0.1088 | -0.0335 | 0.2153 | 0.0013 | 0.4562 |
| Q37 | To curb antimicrobial resistance, regular consultations or ward rounds with a clinical microbiologist or infectious disease physician are useful | 0.7046 | 0.07870 | 0.0061 | 0.0190 | 0.1881 | 0.0781 | 0.4555 |
|  |  | Factor 1 | Factor 2 | Factor 3 | Factor 4 | Factor 5 | Factor 6 |  |
| **Q38** | To curb antimicrobial resistance, doctors need to have timely access to microbiological test results to guide antibiotic therapy | 0.7197 | 0.08350 | 0.2786 | 0.0985 | 0.0670 | 0.0820 | 0.3765 |
| **Q39** | Up-to-date information on hospital antimicrobial resistance patterns is important for developing hospital antibiotic guidelines | 0.7374 | 0.08540 | 0.3223 | 0.1088 | 0.1739 | 0.0348 | 0.3018 |
| **Q40** | Effective infection prevention and control in the hospital reduces antimicrobial resistance | 0.7067 | 0.16900 | 0.2876 | 0.0950 | 0.1798 | 0.0533 | 0.3452 |
| **Eigenvalues** |  | **4.39** | **3.19** | **2.82** | **2.78** | **2.65** | **2.17** |  |
| **% of variance explained** |  | **11.56%** | **8.40%** | **7.43%** | **7.32%** | **6.98%** | **5.72%** | **Overall**<br>**47.4%** |

Table shows the results of the exploratory factor analysis (principal axis factoring) with orthogonal varimax rotation of the six-factor solution using the factor, pcf command in Stata.
Rotated factor loadings: a measure of how much each item contributes to the factor. Loadings close to -1 or 1 indicate that the factor strongly affects the item and loadings close to 0 indicate that the factor has a weak effect on the item.
Uniqueness: shows the proportion of the item's variance that is not explained by the factors.
Item #9 and 10 were excluded from the analysis as explained in Results.

**Table 3.** The latent factors of antibiotic prescribing

| Factor | Factor label | Number of items | Original item # | Loadings range | Reliability<br>(Cronbach's $\alpha$) |
| --- | --- | --- | --- | --- | --- |
| 1 | Awareness of AMS activities | 8 | 33-40 | 0.5202, 0.7374 | 0.8734 |
| 2 | Awareness of AMS purposes | 5 | 28-32 | 0.6224, 0.7744 | 0.8334 |
| 3 | Views regarding rational antibiotic prescribing | 5 | 11, 17-20 | 0.3499, 0.7362 | 0.6961 |
| 4 | Confidence in antibiotic prescribing decisions | 8 | 12, 16, 21, 22, 24-27 | 0.4031, 0.7092 | 0.6997 |
| 5 | Perception of AMR as a significant problem | 6 | 1-6 | 0.4766, 0.5742 | 0.6967 |
| 6 | Immediate actions to contain AMR | 6 | 7, 8, 13-15, 23 | 0.3617, 0.6368 | 0.5695 |

Abbreviations: AMR, antimicrobial resistance; AMS, antibiotic stewardship. Item #9 and 10 were excluded from the analysis, as explained in Results. The full table is included in Table S2.

### Description of respondent responses

Figure 1 and **Table S3** summarise the responses to all 40 items.

**Figure 1.**
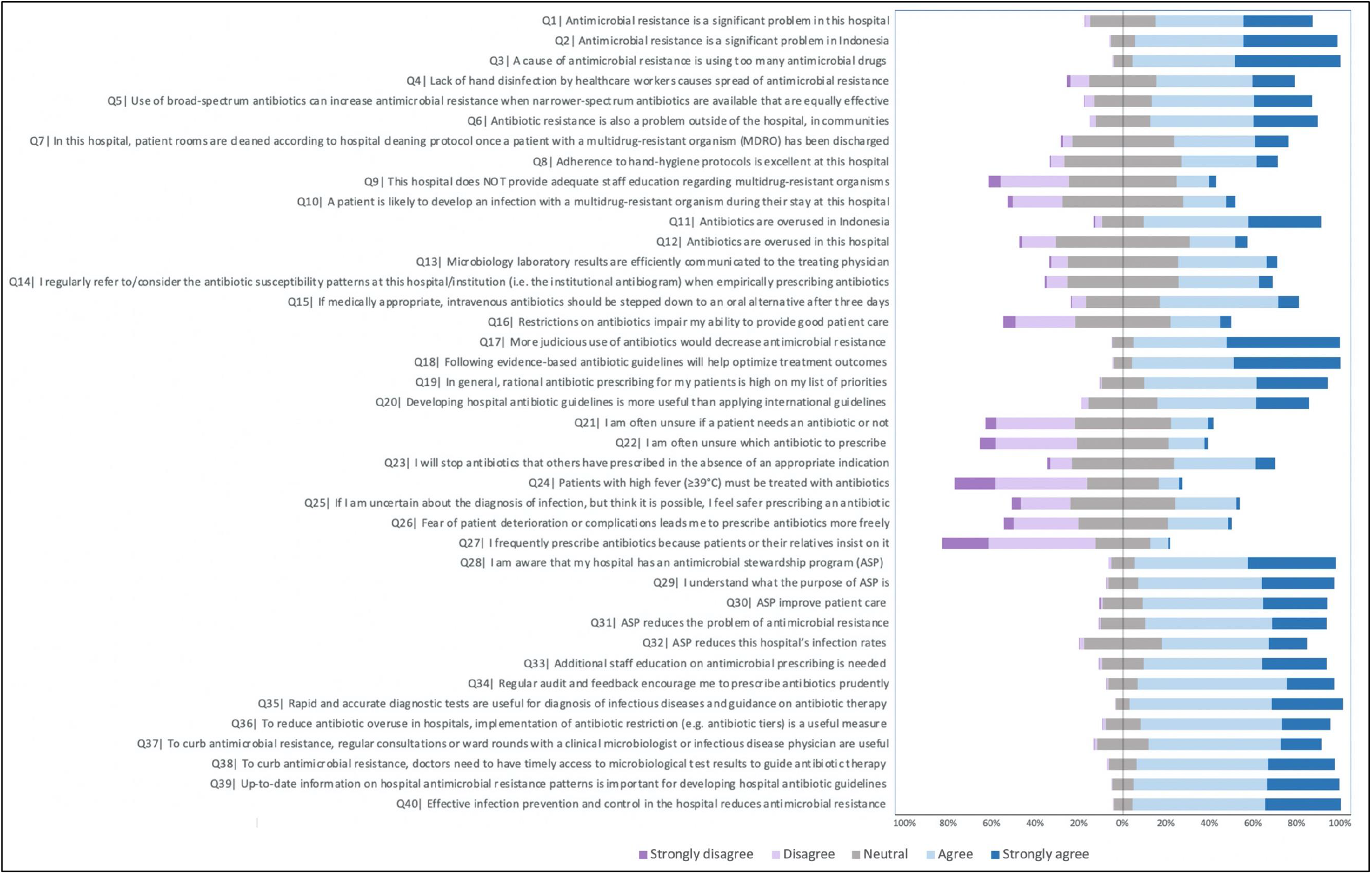
Five-point Likert scale responses for the 40-item questionnaire Acronyms: ASP, antibiotic stewardship program. Data (n, %) are summarized in Table S3.

#### Scope of the AMR problem and key contributors

Most respondents agreed that AMR is an important problem in Indonesia (93.8%; 944/1006) [Q2]; in communities outside of the hospital (83.6%; 838/1003) [Q6]; and at their hospital (80.4%; 808/1005) [Q1], with 30.9% (311/1005) agreeing that patients are likely to develop an infection with a multidrug-resistant infection [Q10]. Most acknowledged as key contributing factors: overuse of antimicrobial drugs (95.1%; 954/1003) [Q3], lack of hand hygiene (71.1%; 715/1005) [Q4], use of broad-spectrum antibiotics (80.5%; 808/1004) [Q5]. Current infection and prevention control (IPC) practices at their hospital were regarded as suboptimal: 64.8% (651/1004) thought that patient rooms are cleaned according to hospital protocol after discharge of a patient with a multidrug-resistant organism [Q7]; 56.9% (570/1002) thought adherence to hand-hygiene protocols to be excellent [Q8]; and 22.5% (226/1005) felt that their hospital does not provide adequate staff education regarding multidrug-resistant organisms [Q9].

#### Antibiotic prescribing practices

Whereas most respondents (85.7%; 861/1005) agreed that antibiotics are overused in Indonesia (Q11), only 35.5% (357/1005) acknowledged this to be case at their hospital [Q12]. Most agreed that more judicious antimicrobial prescribing practices would decrease AMR (94.8%; 953/1005) [Q17] and that following evidence-based antibiotic guidelines will help optimize treatment outcomes (95.3%; 958/1006) [Q18]. Most gave high priority to rational antibiotic prescribing to their patients (88.8%; 892/1005) [Q19], and considered developing hospital antibiotic guidelines more useful than applying international guidelines (78.4%; 787/1004) [Q20]. Nearly a quarter of respondents indicated to be often unsure if a patient needs an antibiotic or not (23.6%; 237/1006) [Q21] or which antibiotic to prescribe (21.3%; 215/1006) [Q22]. A small but considerable fraction expressed lack of confidence in prescribing decisions, i.e. 12.2% (123/1005) prescribed in patients with just a high fever (≥39ºC) [Q24], 36.6% (368/1005) when they felt uncertain about the diagnosis of infection [Q25], 35.0% (352/1006) prescribed more freely because of fear of clinical failure [Q26], and 9.8% (98/1005) frequently prescribed antibiotics because patients or their relatives insist [Q27]. Just more than half of the respondents reported that microbiology laboratory results are efficiently communicated to the treating physician (57.3%; 576/1005) [Q13], considered the hospital antibiogram when empirically prescribing antibiotics (54.5%; 548/1005) [Q14], and would stop antibiotics that others have prescribed in the absence of an appropriate indication (57.0%; 571/1002) [Q23]. Most (72.7%; 731/1006) agreed that, if medically appropriate, intravenous antibiotics should be stepped down to an oral alternative after three days [Q15]. Notably, 33.6% (338/1006) felt that restrictions on antibiotics impaired their ability to provide good patient care [Q16].

#### Antibiotic stewardship

When asked about AMS in general, most respondents were aware that their hospital had an AMS programme (93.1%; 937/1006) [Q28], they reported they understood its purpose (92.1%; 927/1006) [Q29], and they agreed that AMS can improve patient care (88.6%; 891/1006) [Q30], reduce AMR (88.3%; 887/1005) [Q31] and hospital-acquired infections (76.7%; 770/1004) [Q32]. When asked about the usefulness of specific AMS activities, most respondents acknowledged that additional education on antibiotic prescribing was needed (88.4%; 888/1005) [Q33], regular audit and feedback encouraged them to prescribe antibiotics prudently (92.2%; 928/1006) [Q34], rapid and accurate diagnostic tests are useful for diagnosis of infectious diseases and guidance on antibiotic therapy (96.5%; 971/1006) [Q35], implementation of antibiotic restriction (e.g. antibiotic tiers) can reduce antibiotic overuse in hospitals (90.4%; 910/1007) [Q36], regular consultations or ward rounds with a clinical microbiologist or infectious disease physician can curb AMR (85.4%; 859/1006) [Q37], timely access to microbiological test results is needed to guide antibiotic therapy (92.4%; 930/1006) [Q38], and IPC in the hospital can reduce AMR (95.3%; 959/1006) [Q40].

### Physician subgroup analysis

**Table 4** summarizes the results of the subgroup analysis.

**Table 4.** Physician subgroup analysis

|  | Factor 1 |  |  | Factor 2 |  |  | Factor 3 |  |  | Factor 4 |  |  | Factor 5 |  |  | Factor 6 |  |  |
| --- | --- | --- | --- | --- | --- | --- | --- | --- | --- | --- | --- | --- | --- | --- | --- | --- | --- | --- |
|  | Coeff | 95% CI | P | Coeff | 95% CI | P | Coeff | 95% CI | P | Coeff | 95% CI | P | Coeff | 95% CI | P | Coeff | 95% CI | P |
| **Hospital** |  |  |  |  |  |  |  |  |  |  |  |  |  |  |  |  |  |  |
| **05** | Ref |  |  | Ref |  |  | Ref |  |  | Ref |  |  | Ref |  |  | Ref |  |  |
| **01** | 0.194 | -0.089, 0.477 | 0.179 | **-0.422** | **-0.701, -0.142** | **0.003** | 0.244 | -0.039, 0.527 | 0.091 | -0.167 | -0.451, 0.117 | 0.248 | **-0.430** | **-0.716, -0.145** | **0.003** | **-0.504** | **-0.778, -0.23** | **0.000** |
| **02** | 0.064 | -0.13, 0.258 | 0.518 | **-0.352** | **-0.543, -0.161** | **0.000** | 0.134 | -0.06, 0.328 | 0.174 | -0.180 | -0.375, 0.014 | 0.069 | -0.095 | -0.29, 0.101 | 0.342 | -0.087 | -0.274, 0.101 | 0.366 |
| **03** | 0.164 | -0.116, 0.444 | 0.252 | -0.066 | -0.343, 0.211 | 0.641 | **0.387** | **0.107, 0.668** | **0.007** | 0.131 | -0.151, 0.412 | 0.363 | -0.044 | -0.327, 0.238 | 0.758 | 0.092 | -0.18, 0.364 | 0.507 |
| **04** | 0.117 | -0.218, 0.453 | 0.494 | 0.116 | -0.215, 0.448 | 0.492 | 0.096 | -0.239, 0.432 | 0.573 | 0.249 | -0.088, 0.586 | 0.148 | -0.081 | -0.419, 0.257 | 0.638 | **0.361** | **0.035, 0.686** | **0.030** |
| **06** | 0.196 | -0.153, 0.544 | 0.271 | 0.106 | -0.239, 0.45 | 0.548 | **0.419** | **0.07, 0.768** | **0.019** | 0.013 | -0.338, 0.363 | 0.944 | **-0.432** | **-0.783, -0.08** | **0.016** | **0.479** | **0.141, 0.817** | **0.005** |
| **Departments** |  |  |  |  |  |  |  |  |  |  |  |  |  |  |  |  |  |  |
| **Surgical** | Ref |  |  | Ref |  |  | Ref |  |  | Ref |  |  | Ref |  |  | Ref |  |  |
| **Medical** | **0.239** | **0.073, 0.405** | **0.005** | 0.150 | -0.014, 0.314 | 0.073 | -0.120 | -0.286, 0.046 | 0.156 | **0.307** | **0.140, 0.473** | **0.000** | **0.221** | **0.054, 0.389** | **0.009** | **-0.398** | **-0.559, -0.237** | **0.000** |
| **Acute** | **-0.275** | **-0.488, -0.062** | **0.011** | **-0.214** | **-0.425, -0.004** | **0.046** | **-0.273** | **-0.486, -0.06** | **0.012** | 0.169 | -0.045, 0.382 | 0.122 | 0.124 | -0.091, 0.338 | 0.258 | **0.242** | **0.036, 0.449** | **0.021** |
| **Other** | -0.081 | -0.253, 0.092 | 0.358 | -0.027 | -0.197, 0.144 | 0.760 | -0.086 | -0.259, 0.086 | 0.326 | 0.046 | -0.127, 0.219 | 0.603 | 0.119 | -0.055, 0.292 | 0.181 | **-0.302** | **-0.469, -0.135** | **0.000** |
| **Medical** | Ref |  |  | Ref |  |  | Ref |  |  | Ref |  |  | Ref |  |  | Ref |  |  |
| **Acute** | **-0.514** | **-0.745, -0.283** | **0.000** | **-0.364** | **-0.593, -0.136** | **0.002** | -0.153 | -0.384, 0.078 | 0.194 | -0.138 | -0.370, 0.094 | 0.243 | -0.098 | -0.330, 0.135 | 0.411 | **0.640** | **0.417, 0.864** | **0.000** |
| **Other** | **-0.320** | **-0.507, -0.133** | **0.001** | -0.177 | -0.361, 0.0083 | 0.061 | 0.034 | -0.154, 0.221 | 0.724 | **-0.261** | **-0.449, 0.073** | **0.006** | -0.103 | -0.291, 0.086 | 0.285 | 0.096 | -0.085, 0.277 | 0.298 |
| **Medical Hierarchy** |  |  |  |  |  |  |  |  |  |  |  |  |  |  |  |  |  |  |
| **Consultant** | Ref |  |  | Ref |  |  | Ref |  |  | Ref |  |  | Ref |  |  | Ref |  |  |
| **Intern** | **-0.744** | **-1.374, -0.115** | **0.020** | **0.637** | **0.015, 1.259** | **0.045** | 0.443 | -0.187, 1.074 | 0.168 | 0.067 | -0.565, 0.699 | 0.836 | 0.495 | -0.14, 1.129 | 0.126 | 0.003 | -0.607, 0.613 | 0.993 |
| **GP** | 0.168 | -0.096, 0.432 | 0.212 | 0.153 | -0.108, 0.413 | 0.251 | **-0.397** | **-0.661, -0.133** | **0.003** | -0.169 | -0.434, 0.095 | 0.210 | -0.011 | -0.276, 0.255 | 0.938 | -0.024 | -0.279, 0.232 | 0.857 |
| **Resident** | -0.091 | -0.287, 0.105 | 0.363 | **0.297** | **0.104, 0.490** | **0.003** | 0.004 | -0.192, 0.199 | 0.971 | **-0.271** | **-0.468, -0.075** | **0.007** | 0.058 | -0.139, 0.255 | 0.563 | **-0.202** | **-0.391, -0.012** | **0.037** |
| **Other** | -0.013 | -0.491, 0.466 | 0.958 | 0.148 | -0.325, 0.620 | 0.541 | 0.252 | -0.227, 0.731 | 0.303 | -0.172 | -0.653, 0.308 | 0.482 | -0.013 | -0.495, 0.469 | 0.959 | 0.304 | -0.159, 0.768 | 0.198 |
| **Intern** | Ref |  |  | Ref |  |  | Ref |  |  | Ref |  |  | Ref |  |  | Ref |  |  |
| **GP** | **0.912** | **0.245, 1.579** | **0.007** | -0.485 | -1.144, 0.175 | 0.150 | -0.840 | -1.508, -0.172 | 0.014 | -0.236 | -0.906, 0.434 | 0.489 | -0.505 | -1.18, 0.167 | 0.141 | -0.0263 | -0.673, 0.620 | 0.936 |
| **Resident** | **0.653** | **0.0458, 1.261** | **0.035** | -0.340 | -0.941, -0.260 | 0.267 | -0.440 | -1.048, 0.169 | 0.157 | -0.338 | -0.948, 0.272 | 0.278 | -0.436 | -1.049, 0.176 | 0.162 | -0.205 | -0.793, 0.384 | 0.496 |
| **Other** | 0.732 | -0.457, 1.509 | 0.065 | -0.490 | -1.258, 0.279 | 0.212 | -0.191 | -0.970, 0.587 | 0.630 | -0.239 | -1.020, 0.542 | 0.548 | -0.507 | -1.291, 0.276 | 0.204 | 0.302 | -0.452, 1.055 | 0.433 |
| **Resident** | Ref |  |  | Ref |  |  | Ref |  |  | Ref |  |  | Ref |  |  | Ref |  |  |
| **GP** | 0.259 | -0.037, 0.555 | 0.086 | -0.145 | -0.437, 0.148 | 0.333 | -0.400 | -0.696, -0.104 | 0.008 | 0.102 | -0.196, 0.399 | 0.503 | -0.069 | -0.367, 0.229 | 0.651 | 0.178 | -0.108, 0.465 | 0.223 |
| **Other** | 0.0781 | -0.417, 0.574 | 0.757 | -0.150 | -0.639, 0.340 | 0.550 | 0.248 | -0.248, 0.744 | 0.326 | 0.099 | -0.399, 0.596 | 0.697 | -0.710 | -0.570, 0.428 | 0.780 | **0.506** | **-0.163, 1.29** | **0.039** |
| **Years in current profession** |  |  |  |  |  |  |  |  |  |  |  |  |  |  |  |  |  |  |
| **0-5** | Ref |  |  | Ref |  |  | Ref |  |  | Ref |  |  | Ref |  |  | Ref |  |  |
| **6-10** | 0.089 | -0.122, 0.299 | 0.410 | 0.225 | **0.017, 0.434** | **0.034** | -0.089 | -0.300, 0.122 | 0.410 | 0.094 | -0.118, 0.305 | 0.385 | **0.290** | **0.077, 0.502** | **0.008** | 0.053 | -0.151, 0.258 | 0.608 |
| **11-15** | 0.087 | -0.185, 0.358 | 0.532 | -0.015 | -0.283, 0.254 | 0.915 | -0.066 | -0.338, 0.207 | 0.637 | 0.271 | -0.002, 0.544 | 0.052 | 0.156 | -0.118, 0.43 | 0.265 | -0.095 | -0.358, 0.169 | 0.481 |
|  | Coeff | 95% CI | P | Coeff | 95% CI | P | Coeff | 95% CI | P | Coeff | 95% CI | P | Coeff | 95% CI | P | Coeff | 95% CI | P |
| >15 | 0.117 | -0.118, 0.353 | 0.328 | 0.194 | -0.039, 0.427 | 0.102 | -0.044 | -0.279, 0.192 | 0.715 | -0.057 | -0.293, 0.179 | 0.637 | **0.333** | **0.096, 0.57** | **0.006** | **0.366** | **0.138, 0.594** | **0.002** |
| **Public sector\*** | -0.064 | -0.4, 0.273 | 0.710 | 0.264 | -0.207, 0.736 | 0.272 | -0.238 | -0.575, 0.100 | 0.167 | 0.256 | -0.835, 0.595 | 0.140 | 0.192 | -0.148, 0.533 | 0.268 | 0.362 | 0.273, 0.996 | 0.264 |
| **Tertiary level\*** | -0.093 | -0.42, 0.234 | 0.577 | -0.289 | -0.767, 0.190 | 0.237 | -0.047 | -0.375, 0.281 | 0.779 | -0.317 | -0.647, 0.012 | 0.059 | 0.052 | -0.279, 0.383 | 0.758 | -0.407 | -1.065, 0.25 | 0.225 |

Abbreviations: GP, general practitioner.
The table summarizes the results of the multivariable mixed-effect linear regression models to assess the associations between independent variables and each of the factor scores (dependent variable), while adjusting for the possible interdependence of observations clustered within hospitals as well as confounders (sex, AMS training).
\* This model did not include hospital as an independent variable due to collinearity.

#### Hospitals

Statistically significant differences were identified between hospitals for awareness of AMS purposes (factor 2), views regarding rational antibiotic prescribing (factor 3), perception of AMR as a significant problem (factor 5) and immediate actions to contain AMR (factor 6), but not for awareness of AMS activities (factor 1) and confidence in antibiotic prescribing decisions (factor 4). None of the factor scores differed between prescribers in public versus private, or secondary versus tertiary hospitals.

#### Professional hierarchy

For awareness of AMS activities (factor 1), consultants, GPs and residents scored higher than interns. For awareness of AMS purposes (factor 2), consultants scored lower than interns and residents. For views regarding rational antibiotic prescribing (factor 3), consultants scored higher than GPs. For confidence in antibiotic prescribing decisions (factor 4), consultants scored lower than residents, whereas for immediate actions to contain AMR (factor 6), consultants scored higher than residents. No differences were identified for perception of AMR as a significant problem (factor 5).

#### Departments

For awareness of AMS activities (factor 1) and purpose (factor 2), physicians in surgery and medicine scored higher than the acute specialties, whereas for awareness of AMS activities (factor 1), medicine scored higher than surgery. For views regarding rational antibiotic prescribing (factor 3), surgery scored higher than acute specialties. For confidence in antibiotic prescribing decisions (factor 4), medicine scored lower than surgery and other specialties. For perception of AMR as a significant problem (factor 5), surgery scored lower than medicine. For immediate actions to contain AMR (factor 6), surgery scored higher than medicine and other specialties, and the acute specialties scored higher than surgery and medicine.

#### Work experience

Physicians with little (0-5 years) work experience scored lower than more experienced colleagues, for awareness of AMS purpose (factor 2); for perception of AMR as a significant problem (factor 5); and for immediate actions to contain AMR (factor 6). No differences were identified for factors 1, 3 and 4.

## Discussion

This survey assessed the perceptions, views and practices regarding AMR, antibiotic prescribing and AMS among over 1000 physicians in Indonesian hospitals. Through an exploratory factor analysis we identified six distinct constructs in the dataset, i.e., 1) awareness of AMS activities; 2) awareness of AMS purposes; 3) views regarding rational antibiotic prescribing; 4) confidence in antibiotic prescribing decisions; 5) perception of AMR as a significant problem; and 6) immediate actions to contain AMR. The survey findings outline a series of dynamics around AMR and AMS in the Indonesian context. Spanning issues around *visibility* (diagnostics)^26^, *awareness* (education)^27^ and *institutional form* (governance)^28^, the survey results tease out many of the core issues illustrated in other settings^4,5^, but in turn, illustrate a specific mix of variables at play, i.e. uncertainty, risk, and lack of sense of responsibility. For instance, only about one-third of physicians recognised that antibiotic overuse was an issue at their own hospital, many physicians were hesitant to stop antibiotics that others had prescribed in the absence of an appropriate indication, and felt that antibiotic restrictions impaired their ability to provide good patient care. Lack of confidence in prescribing decisions and defensive prescribing were common due to diagnostic uncertainty, fear of patient deterioration or complications, or because patients or their relatives insisted. The study findings expand on our recently published paper on the patterns and quality indicators of antibiotic prescribing in the same hospitals, which identified several priority areas for stewardship^17^.

The most significant factor in guiding the future agenda in Indonesia around effective AMS implementation, is the perceived “externality of AMR” as a problem^29^. That is, physicians acknowledge its significance but do not take ownership or responsibility, thus reflecting a production of AMR as an externality, e.g. a result of irrational use elsewhere in communities or other hospitals. The lack of systematic surveillance of AMR and antibiotic use and the underutilisation of bacterial cultures, recognised by many of the respondents, also reproduces the perception of AMR as a “problem of elsewhere”. This feeds a lack of engaging with AMS, as it is not recognised as a value-add for an already stretched institutional context, and in turn provides the context for continued defensive prescribing “to be on the safe side”. Moreover, defensive prescribing practices somewhat offset (in the short term) problems around room cleaning, hand hygiene and staff education. In this way, AMR as an externality and the vulnerabilities of the institution, offer an environment conducive to the ongoing over-use of antimicrobials^30,31^. The higher incidence of hospital-acquired infections in LMIC than in high-income countries could further promote defensive prescribing as a way to compensate for substandard IPC practices^32^. All in all, this supports the notion that physicians tend to prioritise managing immediate clinical risks, reputation and concordance with peer practice, vis-à-vis the long-term population consequences of AMR^33^. Work experience and medical hierarchy were found to influence the awareness of AMS purpose, AMR as a significant problem, and immediate actions to contain AMR. Interestingly, compared with junior physicians, specialists/consultants expressed lower confidence to make antibiotic decisions in uncertain situations while showing higher confidence in actions to contain AMR. Possible explanations include that specialists/consultants have a better recognition of the “unknowns” (e.g. lack of data on bacterial susceptibility patterns) and that they bear final patient responsibility, introducing the fear of losing a patient or legal consequences^34^, whereas taking actions to curb AMR can be a remedy to compensate their fear. Conversely, residents’ higher confidence in antibiotic prescribing may also relate to their contemporary medical training, which includes AMS, as opposed to late-career physicians^35^. GPs had low scores on views regarding rational antibiotic prescribing compared with consultants which could reflect the GPs’ limited responsibility in the antibiotic decision-making hierarchy, possibly leading to a lack of positive attitude towards guidelines and preference for complying with them^36,37^.

The acute specialties (including emergency, ICU and anaesthesiology) had lower awareness of AMS activities and purposes, compared with surgery or medicine, but scored higher for immediate actions to contain AMR. Compared with surgeons, physicians in medicine had a greater awareness of AMS activities and recognition of AMR as a significant problem, but they had lower confidence in antibiotic prescribing decisions and immediate actions to contain AMR. These observations are in line with a UK study that found that emergency physicians experienced pressure for immediate action out of fear of losing a patient and a lack of ownership of antibiotic decision-making due to the patient transitioning to inpatient care, that medical doctors adopted a more policy-informed, interdisciplinary approach, and that senior surgeons left complex antibiotic decisions to junior staff, resulting in potential defensive and inappropriate antibiotic use^38^. Variations in the social norms, values and behaviours between specialties should inform what is the best approach to antibiotic decision-making.

Our study had several strengths and weaknesses. First, based on our self-developed questionnaire, we were able to identify a relevant set of attributes through a factor analysis optimization process, with adequate content, face and construct validity and internal reliability. In the absence of adequately validated instruments regarding AMR and AMS^39^, this study adds important value to the field, with particular relevance and applicability for LMIC. Nonetheless, further questionnaire validation steps (such as criterion-related validity) are necessary to achieve a fully valid and reliable instrument. Second, the study had a large, varied respondent sample and high response rate. However, non-participation and the convenient hospital sample could have introduced selection bias, and the data are not necessarily representative for Jakarta or Indonesia at large. The authenticity of the answers was maximised by protecting the respondents’ identities, although reliance on self-report has potential for social desirability bias. Third, factor analysis is based on using a “heuristic”, which leaves room to more than one interpretation of the same data and cannot identify causality.

## Conclusion

AMS implementation in Indonesian hospitals is likely highly dependent on institutional, contextual and diagnostic vulnerabilities. These may result in the problem of AMR being externalised, instead of recognised as a local hospital problem. Current AMS strategies may be insufficiently successful in promoting prudent antibiotic use, due to lack of systematic engagement with and feedback to prescribers, aimed at building confidence in antibiotic decision-making and ownership of the AMR problem. Appropriate recognition of the contextual and social determinants of antibiotic prescribing decision-making, including hospital factors, dynamics in medical hierarchy and experience, among others, will be critical to design context-specific AMS interventions that are adopted by healthcare professionals and successfully influence behaviours^12^.

## Data Availability

De-identified data are available upon reasonable request via the corresponding author, after written permission has been obtained from the lead investigators.

## Acknowledgements

The authors are grateful to the management, research/medical committees and clinicians of the participating hospitals for their support to the study.

## EXPLAIN study group

Ralalicia Limato, Erni J. Nelwan, Manzilina Mudia, Monik Alamanda, Helio Guterres, Enty Enty, Elfrida R. Manurung, Ifael Y. Mauleti, Maria Mayasari, Iman Firmansyah, May Hizrani, Roswin Djafar, Raph L. Hamers, Anis Karuniawati, Prof Taralan Tambunan, Prof Amin Soebandrio, Decy Subekti, Iqbal Elyazar, Mutia Rahardjani, Fitria Wulandari, Prof Reinout van Crevel, H. Rogier van Doorn, Vu Thi Lan Huong, Nga Do Ti Thuy, Sonia Lewycka, Prof Alex Broom

## Author contributions

EJN and RLH conceived the idea for the study and are the principal investigators. RLH and RL obtained the funding. RL, EJN, HVTL and RLH designed the study protocol and developed the study instrument. MM, ERM, IYM, MM, IF, and RD collected and verified the data, overseen by RL. RL, MM, MA created and curated the database. RLH and MA performed the analysis and had full access to all study data. RL, MA, and RLH drafted the paper, with critical inputs from EJN, HVTL, HRvD and AB. All authors critically revised the manuscript and gave approval for the final version to be published.

## Appendix: Survey questionnaire

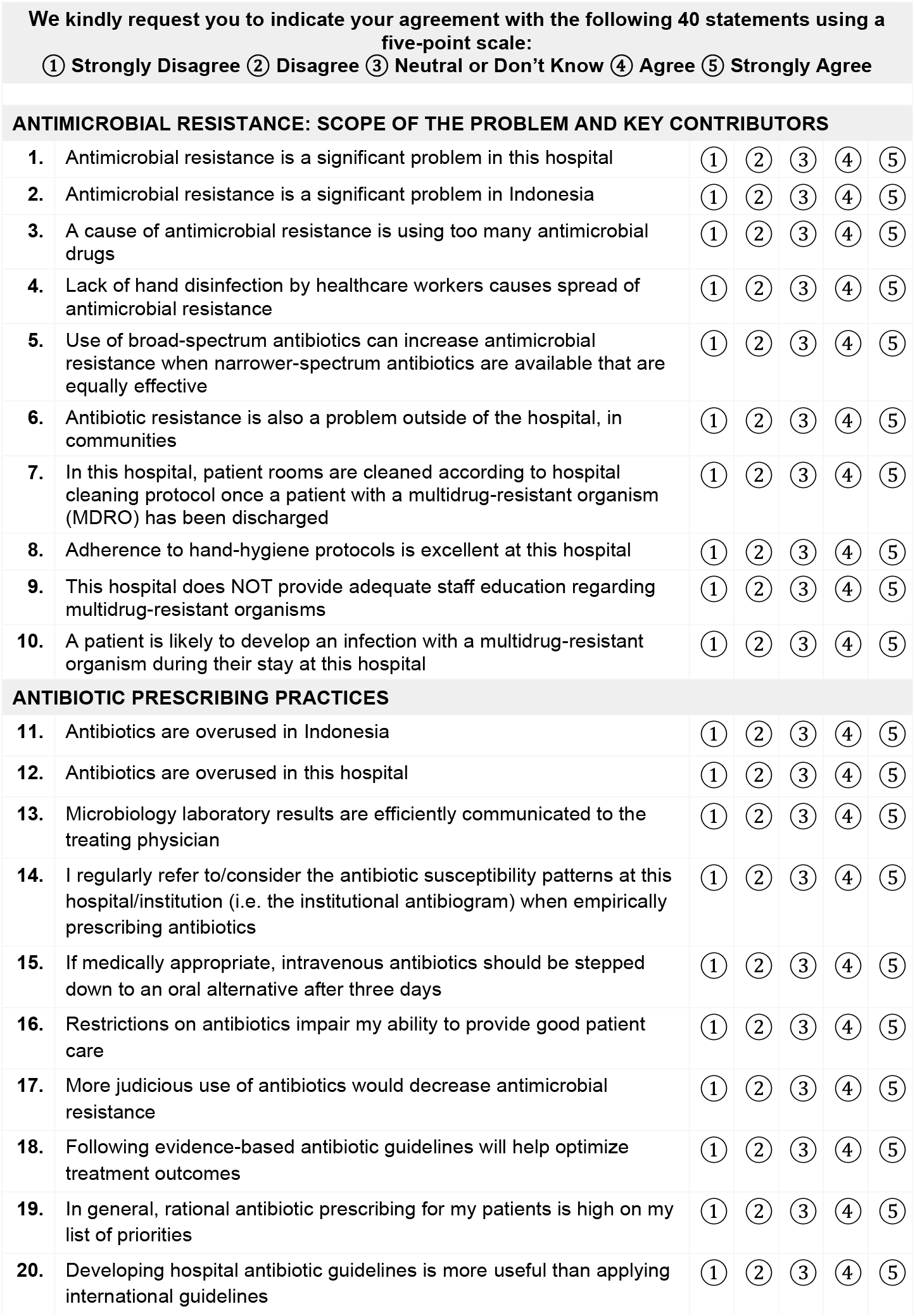
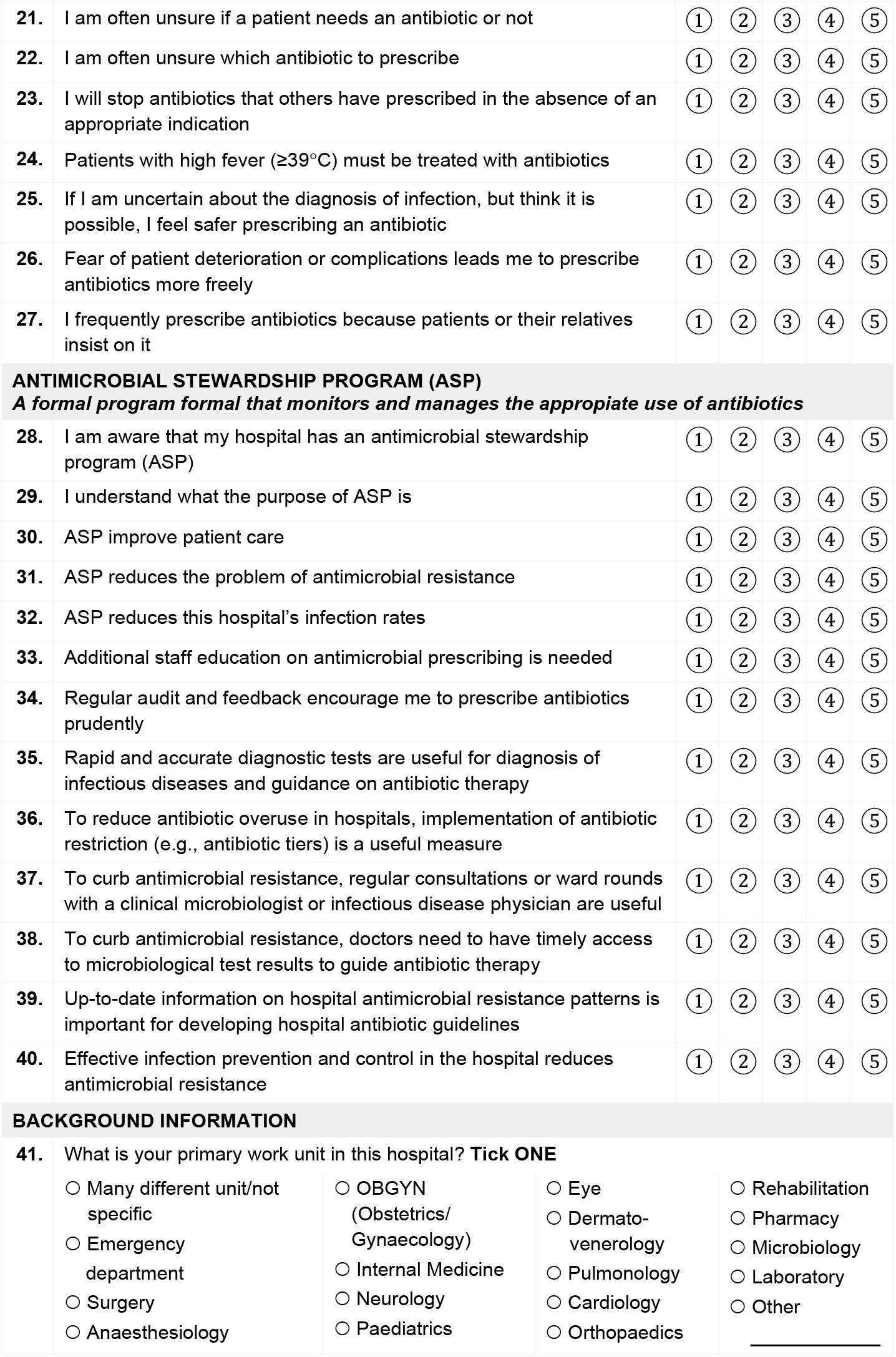
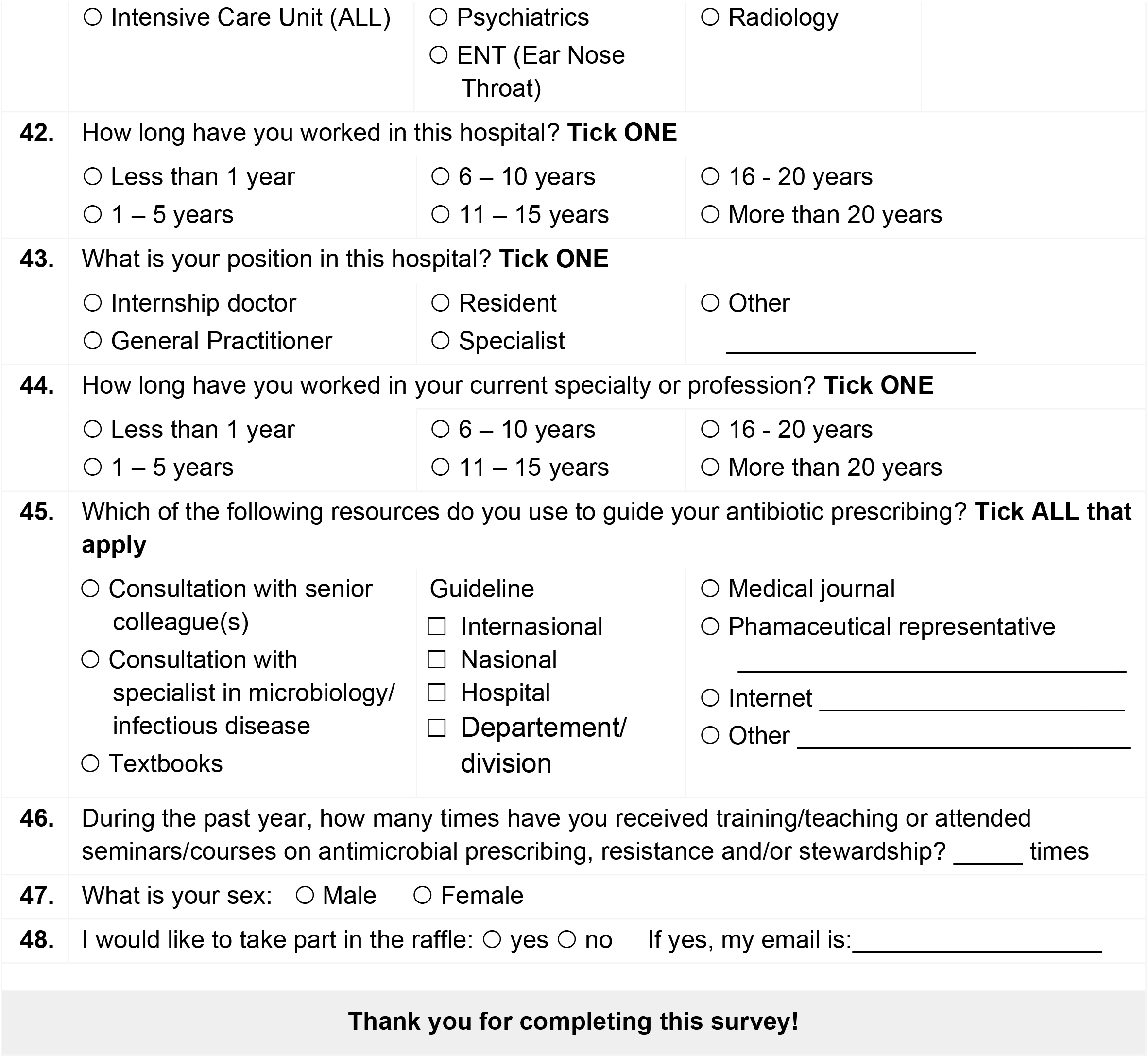

ANTIMICROBIAL RESISTANCE: SCOPE OF THE PROBLEM AND KEY CONTRIBUTORS
|  |  |  |  |  |  |  |
| --- | --- | --- | --- | --- | --- | --- |
| 1. | Antimicrobial resistance is a significant problem in this hospital | ① | ② | ③ | ④ | ⑤ |
| 2. | Antimicrobial resistance is a significant problem in Indonesia | ① | ② | ③ | ④ | ⑤ |
| 3. | A cause of antimicrobial resistance is using too many antimicrobial drugs | ① | ② | ③ | ④ | ⑤ |
| 4. | Lack of hand disinfection by healthcare workers causes spread of antimicrobial resistance | ① | ② | ③ | ④ | ⑤ |
| 5. | Use of broad-spectrum antibiotics can increase antimicrobial resistance when narrower-spectrum antibiotics are available that are equally effective | ① | ② | ③ | ④ | ⑤ |
| 6. | Antibiotic resistance is also a problem outside of the hospital, in communities | ① | ② | ③ | ④ | ⑤ |
| 7. | In this hospital, patient rooms are cleaned according to hospital cleaning protocol once a patient with a multidrug-resistant organism (MDRO) has been discharged | ① | ② | ③ | ④ | ⑤ |
| 8. | Adherence to hand-hygiene protocols is excellent at this hospital | ① | ② | ③ | ④ | ⑤ |
| 9. | This hospital does NOT provide adequate staff education regarding multidrug-resistant organisms | ① | ② | ③ | ④ | ⑤ |
| 10. | A patient is likely to develop an infection with a multidrug-resistant organism during their stay at this hospital | ① | ② | ③ | ④ | ⑤ |
| 11. | Antibiotics are overused in Indonesia | ① | ② | ③ | ④ | ⑤ |
| 12. | Antibiotics are overused in this hospital | ① | ② | ③ | ④ | ⑤ |
| 13. | Microbiology laboratory results are efficiently communicated to the treating physician | ① | ② | ③ | ④ | ⑤ |
| 14. | I regularly refer to/consider the antibiotic susceptibility patterns at this hospital/institution (i.e. the institutional antibiogram) when empirically prescribing antibiotics | ① | ② | ③ | ④ | ⑤ |
| 15. | If medically appropriate, intravenous antibiotics should be stepped down to an oral alternative after three days | ① | ② | ③ | ④ | ⑤ |
| 16. | Restrictions on antibiotics impair my ability to provide good patient care | ① | ② | ③ | ④ | ⑤ |
| 17. | More judicious use of antibiotics would decrease antimicrobial resistance | ① | ② | ③ | ④ | ⑤ |
| 18. | Following evidence-based antibiotic guidelines will help optimize treatment outcomes | ① | ② | ③ | ④ | ⑤ |
| 19. | In general, rational antibiotic prescribing for my patients is high on my list of priorities | ① | ② | ③ | ④ | ⑤ |
| 20. | Developing hospital antibiotic guidelines is more useful than applying international guidelines | ① | ② | ③ | ④ | ⑤ |
| 21. | I am often unsure if a patient needs an antibiotic or not | ① | ② | ③ | ④ | ⑤ |
| 22. | I am often unsure which antibiotic to prescribe | ① | ② | ③ | ④ | ⑤ |
| 23. | I will stop antibiotics that others have prescribed in the absence of an appropriate indication | ① | ② | ③ | ④ | ⑤ |
| 24. | Patients with high fever ($\geq 39^{\circ}\text{C}$) must be treated with antibiotics | ① | ② | ③ | ④ | ⑤ |
| 25. | If I am uncertain about the diagnosis of infection, but think it is possible, I feel safer prescribing an antibiotic | ① | ② | ③ | ④ | ⑤ |
| 26. | Fear of patient deterioration or complications leads me to prescribe antibiotics more freely | ① | ② | ③ | ④ | ⑤ |
| 27. | I frequently prescribe antibiotics because patients or their relatives insist on it | ① | ② | ③ | ④ | ⑤ |
| 28. | I am aware that my hospital has an antimicrobial stewardship program (ASP) | ① | ② | ③ | ④ | ⑤ |
| 29. | I understand what the purpose of ASP is | ① | ② | ③ | ④ | ⑤ |
| 30. | ASP improve patient care | ① | ② | ③ | ④ | ⑤ |
| 31. | ASP reduces the problem of antimicrobial resistance | ① | ② | ③ | ④ | ⑤ |
| 32. | ASP reduces this hospital's infection rates | ① | ② | ③ | ④ | ⑤ |
| 33. | Additional staff education on antimicrobial prescribing is needed | ① | ② | ③ | ④ | ⑤ |
| 34. | Regular audit and feedback encourage me to prescribe antibiotics prudently | ① | ② | ③ | ④ | ⑤ |
| 35. | Rapid and accurate diagnostic tests are useful for diagnosis of infectious diseases and guidance on antibiotic therapy | ① | ② | ③ | ④ | ⑤ |
| 36. | To reduce antibiotic overuse in hospitals, implementation of antibiotic restriction (e.g., antibiotic tiers) is a useful measure | ① | ② | ③ | ④ | ⑤ |
| 37. | To curb antimicrobial resistance, regular consultations or ward rounds with a clinical microbiologist or infectious disease physician are useful | ① | ② | ③ | ④ | ⑤ |
| 38. | To curb antimicrobial resistance, doctors need to have timely access to microbiological test results to guide antibiotic therapy | ① | ② | ③ | ④ | ⑤ |
| 39. | Up-to-date information on hospital antimicrobial resistance patterns is important for developing hospital antibiotic guidelines | ① | ② | ③ | ④ | ⑤ |
| 40. | Effective infection prevention and control in the hospital reduces antimicrobial resistance | ① | ② | ③ | ④ | ⑤ |

BACKGROUND INFORMATION
|  |  |  |  |  |
| --- | --- | --- | --- | --- |
| 41. | What is your primary work unit in this hospital? **Tick ONE** |  |  |  |
|  | ☐ Many different unit/not specific<br>☐ Emergency department<br>☐ Surgery<br>☐ Anaesthesiology | ☐ OBGYN (Obstetrics/ Gynaecology)<br>☐ Internal Medicine<br>☐ Neurology<br>☐ Paediatrics | ☐ Eye<br>☐ Dermato-venerology<br>☐ Pulmonology<br>☐ Cardiology<br>☐ Orthopaedics | ☐ Rehabilitation<br>☐ Pharmacy<br>☐ Microbiology<br>☐ Laboratory<br>☐ Other _____ |

|  |  |  |  |
| --- | --- | --- | --- |
|  | ☐ Intensive Care Unit (ALL) | ☐ Psychiatrics<br>☐ ENT (Ear Nose Throat) | ☐ Radiology |
| **42.** | How long have you worked in this hospital? **Tick ONE** |  |  |
|  | ☐ Less than 1 year<br>☐ 1 – 5 years | ☐ 6 – 10 years<br>☐ 11 – 15 years | ☐ 16 - 20 years<br>☐ More than 20 years |
| **43.** | What is your position in this hospital? **Tick ONE** |  |  |
|  | ☐ Internship doctor<br>☐ General Practitioner | ☐ Resident<br>☐ Specialist | ☐ Other _____ |
| **44.** | How long have you worked in your current specialty or profession? **Tick ONE** |  |  |
|  | ☐ Less than 1 year<br>☐ 1 – 5 years | ☐ 6 – 10 years<br>☐ 11 – 15 years | ☐ 16 - 20 years<br>☐ More than 20 years |
| **45.** | Which of the following resources do you use to guide your antibiotic prescribing? **Tick ALL that apply** |  |  |
|  | ☐ Consultation with senior colleague(s)<br>☐ Consultation with specialist in microbiology/ infectious disease<br>☐ Textbooks | Guideline<br>☐ Internasional<br>☐ Nasional<br>☐ Hospital<br>☐ Departement/ division | ☐ Medical journal<br>☐ Phamaceutical representative<br>_____<br>☐ Internet _____<br>☐ Other _____ |
| **46.** | During the past year, how many times have you received training/teaching or attended seminars/courses on antimicrobial prescribing, resistance and/or stewardship? _____ times |  |  |
| **47.** | What is your sex: ☐ Male ☐ Female |  |  |
| **48.** | I would like to take part in the raffle: ☐ yes ☐ no If yes, my email is: _____ |  |  |
| **Thank you for completing this survey!** |  |  |  |

**Table S1.** Survey response rate, overall and by hospital, department and professional hierarchy

|  | Total physicians invited | Total physicians responded | Response rate (%) | Respondents per physician type |  |  |  |  |
| --- | --- | --- | --- | --- | --- | --- | --- | --- |
|  |  |  |  | Specialist/Consultant | Resident | General practitioner | Intern/Internship doctor | Other |
| **Total** | **1896** | **1007** | **53.1%** | **358 (35.6%)** | **500 (49.7%)** | **113 (11.2%)** | **10 (1.0%)** | **18 (1.8%)** |
| **Hospital <sup>a</sup>** |  |  |  |  |  |  |  |  |
| 01 | 101 (5.3%) | 70 (7.0%) | 69.3% | 39 (10.9%) | 0 (0.0%) | 27 (23.9%) | 0 (0.0%) | 4 (22.2%) |
| 02 | 255 (13.4%) | 155 (15.4%) | 60.8% | 70 (19.6%) | 52 (10.4%) | 26 (23.0%) | 0 (0.0%) | 5 (27.8%) |
| 03 | 143 (7.5%) | 77 (7.7%) | 53.9% | 41 (11.5%) | 0 (0.0%) | 35 (31.0%) | 0 (0.0%) | 1 (5.6%) |
| 04 | 55 (2.9%) | 44 (4.4%) | 80.0% | 29 (8.1%) | 0 (0.0%) | 11 (9.7%) | 0 (0.0%) | 1 (5.6%) |
| 05 | 1240 (65.4%) | 622 (61.8%) | 50.2% | 152 (42.5%) | 448 (89.6%) | 4 (3.5%) | 10 (100.0%) | 5 (27.8%) |
| 06 | 102 (5.4%) | 39 (3.9%) | 38.2% | 27 (7.5%) | 0 (0.0%) | 10 (8.9%) | 0 (0.0%) | 2 (11.1%) |
| **Sector <sup>b</sup>** |  |  |  |  |  |  |  |  |
| Private | 346 | 186 (18.5%) | 53.8% | 107 (29.9%) | 0 (0.0%) | 0 (0.0%) | 72 (63.7%) | 7 (38.9%) |
| Public | 1550 | 821 (81.5%) | 53.0% | 251 (70.1%) | 500 (100.0%) | 41 (36.3%) | 10 (100.0%) | 11 (61.1%) |
| **Level of care <sup>c</sup>** |  |  |  |  |  |  |  |  |
| Secondary | 1499 | 777 (77.2%) | 51.8% | 136 (38.0%) | 0 (0.0%) | 83 (73.5%) | 0 (0.0%) | 8 (44.4%) |
| Tertiary | 397 | 230 (22.8%) | 57.9% | 222 (62.0%) | 500 (100.0%) | 30 (26.6%) | 10 (100.0%) | 10 (55.6%) |
| **Department <sup>d</sup>** |  |  |  |  |  |  |  |  |
| ***Surgery and surgical subspecialties*** | **628** | **371 (36.8)** | **59.1%** | **91 (24.5%)** | **226 (60.9%)** | **1 (0.27%)** | **4 (1.1%)** | **0 (0.0%)** |
| Obstetrics/gynaecology | 229 | 146 (14.5%) | 63.8% | 51 (14.3%) | 91 (18.2%) | 1 (0.88%) | 1 (10.0%) | 0 (0.0%) |
| Surgery | 210 | 122 (12.1%) | 58.1% | 2 (11.1%) | 74 (14.8%) | 0 (0.0%) | 3 (30.0%) | 0 (0.0%) |
| Orthopaedics | 72 | 57 (5.7%) | 44.4% | 25 (7.0%) | 32 (6.4%) | 0 (0.0%) | 0 (0.0%) | 0 (0.0%) |
| Ear-nose-throat | 74 | 32 (3.2%) | 77.0% | 12 (3.4%) | 19 (3.8%) | 0 (0.0%) | 0 (0.0%) | 0 (0.0%) |
| Urology | 43 | 14 (1.4%) | 32.6% | 1 (1.1%) | 10 (2.0%) | 0 (0.0%) | 0 (0.0%) | 0 (0.0%) |
| ***Medicine and medical subspecialties*** | **456** | **232 (23.0%)** | **50.9%** | **115 (49.6%)** | **109 (47.0%)** | **0 (0.0%)** | **3 (1.3%)** | **5 (2.2%)** |
| Internal medicine | 261 | 128 (12.7%) | 49.0% | 59 (16.5%) | 67 (13.4%) | 0 (0.0%) | 0 (0.0%) | 2 (11.1%) |
| Neurology | 74 | 63 (6.3%) | 85.1% | 17 (4.8%) | 40 (8.0%) | 0 (0.0%) | 3 (30.0%) | 3 (16.7%) |
| Pulmonology | 21 | 15 (1.5%) | 71.4% | 15 (4.2%) | 0 (0.0%) | 0 (0.0%) | 0 (0.0%) | 0 (0.0%) |
| Dermato-venerology | 77 | 14 (1.4%) | 18.2% | 12 (0.0%) | 2 (3.4%) | 0 (0.0%) | 0 (0.0%) | 0 (0.0%) |
| Cardiology | 23 | 12 (1.2%) | 52.2% | 12 (3.4%) | 0 (0.0%) | 0 (0.0%) | 0 (0.0%) | 0 (0.0%) |
| ***Acute specialties <sup>e</sup>*** | **96** | **156 (15.5)** | **162.5%** | **28 (17.9%)** | **67 (42.9%)** | **68 (43.6%)** | **2 (1.3%)** | **1 (0.64%)** |
| Anaesthesiology | 96 | 72 (7.2%) | 75.0% | 4 (1.1%) | 66 (13.2%) | 0 (0.0%) | 2 (20.0%) | 0 (0.0%) |
|  |  |  |  | Specialist/Consultant | Resident | General practitioner | Intern/Internship doctor | Other |
| Emergency dept <sup>e</sup> | - | 57 (5.7%) | - | 0 (0.0%) | 0 (0.0%) | 57 (50.4%) | 0 (0.0%) | 0 (0.0%) |
| Intensive Care Unit <sup>e</sup> | - | 27 (2.7%) | - | 14 (3.9%) | 1 (0.2%) | 11 (9.7%) | 0 (0.0%) | 1 (5.6%) |
| **Other departments** | **539** | **244 (24.2)** | **45.2%** | **90 (36.9%)** | **98 (40.2%)** | **24 (9.8%)** | **1 (0.41%)** | **10 (4.1%)** |
| Paediatrics | 202 | 54 (5.4%) | 26.7% | 28 (7.8%) | 25 (5.0%) | 0 (0.0%) | 0 (0.0%) | 0 (0.0%) |
| Ophthalmology | 102 | 39 (3.9%) | 38.2% | 22 (6.2%) | 17 (3.4%) | 0 (0.0%) | 0 (0.0%) | 0 (0.0%) |
| Multiple/unspecified units <sup>e</sup> | - | 33 (3.3%) | - | 5 (1.4%) | 5 (1.0%) | 21 (18.6%) | 1 (10.0%) | 0 (0.0%) |
| Rehabilitation | 70 | 32 (3.2%) | 45.7% | 4 (1.1%) | 28 (5.6%) | 0 (0.0%) | 0 (0.0%) | 0 (0.0%) |
| Psychiatry | 54 | 30 (3.0%) | 55.6% | 11 (3.1%) | 19 (3.8%) | 0 (0.0%) | 0 (0.0%) | 0 (0.0%) |
| Dentist | 49 | 27 (2.7%) | 55.1% | 11 (3.1%) | 1 (0.20%) | 3 (2.7%) | 0 (0.0%) | 10 (55.6%) |
| Other | 39 | 25 (2.5%) | 64.1% | 4 (1.1%) | 1 (0.20%) | 20 (17.7%) | 0 (0.0%) | 0 (0.0%) |
| Microbiology | 4 | 3 (0.30%) | 75.0% | 2 (0.56%) | 1 (0.20%) | 0 (0.0%) | 0 (0.0%) | 0 (0.0%) |
| Laboratory | 19 | 1 (0.10%) | 5.3% | 1 (0.28%) | 0 (0.0%) | 0 (0.0%) | 0 (0.0%) | 0 (0.0%) |
| Missing | - | 4 (0.40%) | - | 2 (0.56%) | 1 (0.20%) | 0 (0.0%) | 0 (0.0%) | 0 (0.0%) |

Data are reported as n (%).
Doctor type was missing for 8 respondents, across <sup>a</sup> hospitals (02 (2), 03 (3), and 05 (3)); <sup>b</sup> sector (public 8, private 0); <sup>c</sup> level of care (secondary 3, tertiary 5); and <sup>d</sup> departments (obstetrics/gynaecology 2; ENT 1; paediatrics 1; multiple units 1; dental/oral surgery 2; missing dept 1).
<sup>e</sup> Invitations to participate in the survey were sent out according to primary work units; the discrepancies reported for the acute and other departments result from the fact that n=177 respondents who worked across multiple units (mostly GPs and internship doctors) indicated a different primary department on their questionnaire.

**Table S2.** The latent factors of antibiotic prescribing (full table)

| Factor | Factor label | Survey questions included in the factor loading | Loadings | Reliability (Cronbach's $\alpha$) |
| --- | --- | --- | --- | --- |
| 1 | Awareness of AMS activities | Q33: Additional staff education on antimicrobial prescribing is needed | 0.5202 | 0.8734 |
|  |  | Q34: Regular audit and feedback encourage me to prescribe antibiotics prudently | 0.6151 |  |
|  |  | Q35: Rapid and accurate diagnostic tests are useful for diagnosis of infectious diseases and guidance on antibiotic therapy | 0.6714 |  |
|  |  | Q36: To reduce antibiotic overuse in hospitals, implementation of antibiotic restriction (e.g. antibiotic tiers) is a useful measure | 0.6428 |  |
|  |  | Q37: To curb antimicrobial resistance, regular consultations or ward rounds with a clinical microbiologist or infectious disease physician are useful | 0.7046 |  |
|  |  | Q38: To curb antimicrobial resistance, doctors need to have timely access to microbiological test results to guide antibiotic therapy | 0.7197 |  |
|  |  | Q39: Up-to-date information on hospital antimicrobial resistance patterns is important for developing hospital antibiotic guidelines | 0.7374 |  |
|  |  | Q40: Effective infection prevention and control in the hospital reduces antimicrobial resistance | 0.7067 |  |
| 2 | Awareness of AMS purposes | Q28: I am aware that my hospital has an antimicrobial stewardship program (ASP) | 0.6224 | 0.8334 |
|  |  | Q29: I understand what the purpose of ASP is | 0.6957 |  |
|  |  | Q30: ASP improve patient care | 0.7744 |  |
|  |  | Q31: ASP reduces the problem of antimicrobial resistance | 0.7532 |  |
|  |  | Q32: ASP reduces this hospital's infection rates | 0.6670 |  |
| 3 | Views regarding rational antibiotic prescribing | Q11: Antibiotics are overused in Indonesia | 0.5138 | 0.6961 |
|  |  | Q17: More judicious use of antibiotics would decrease antimicrobial resistance | 0.7362 |  |
|  |  | Q18: Following evidence-based antibiotic guidelines will help optimize treatment outcomes | 0.6565 |  |
|  |  | Q19: In general, rational antibiotic prescribing for my patients is high on my list of priorities | 0.5246 |  |
|  |  | Q20: Developing hospital antibiotic guidelines is more useful than applying international guidelines | 0.3499 |  |
| 4 | Confidence in antibiotic prescribing decisions | Q12: Antibiotics are overused in this hospital | -0.4550 | 0.6997 |
|  |  | Q16: Restrictions on antibiotics impair my ability to provide good patient care | 0.4031 |  |
|  |  | Q21: I am often unsure if a patient needs an antibiotic or not | 0.5640 |  |
|  |  | Q22: I am often unsure which antibiotic to prescribe | 0.5670 |  |
| | | Q24: Patients with high fever ($\geq 39^{\circ}\text{C}$) must be treated with antibiotics | 0.4794 | |
|  |  | Q25: If I am uncertain about the diagnosis of infection, but think it is possible, I feel safer prescribing an antibiotic | 0.6741 |  |
|  |  | Q26: Fear of patient deterioration or complications leads me to prescribe antibiotics more freely | 0.7092 |  |
|  |  | Q27: I frequently prescribe antibiotics because patients or their relatives insist on it | 0.6318 |  |
| 5 | Perception of AMR as a significant problem | Q1: Antimicrobial resistance is a significant problem in this hospital | 0.5701 | 0.6967 |
|  |  | Q2: Antimicrobial resistance is a significant problem in Indonesia | 0.5742 |  |
|  |  | Q3: A cause of antimicrobial resistance is using too many antimicrobial drugs | 0.5361 |  |
|  |  | Q4: Lack of hand disinfection by healthcare workers causes spread of antimicrobial resistance | 0.5725 |  |
|  |  | Q5: Use of broad-spectrum antibiotics can increase antimicrobial resistance when narrower-spectrum antibiotics are available that are equally effective | 0.5480 |  |
|  |  | Q6: Antibiotic resistance is also a problem outside of the hospital, in communities | 0.4766 |  |
| 6 | Immediate actions to contain AMR | Q7: In this hospital, patient rooms are cleaned according to hospital cleaning protocol once a patient with a multidrug-resistant organism (MDRO) has been discharged | 0.6058 | 0.5695 |
|  |  | Q8: Adherence to hand-hygiene protocols is excellent at this hospital | 0.6368 |  |
|  |  | Q13: Microbiology laboratory results are efficiently communicated to the treating physician | 0.5116 |  |
|  |  | Q14: I regularly refer to/consider the antibiotic susceptibility patterns at this hospital/institution (i.e. the institutional antibiogram) when empirically prescribing antibiotics | 0.6115 |  |
|  |  | Q15: If medically appropriate, intravenous antibiotics should be stepped down to an oral alternative after three days | 0.3617 |  |
|  |  | Q23: I will stop antibiotics that others have prescribed in the absence of an appropriate indication | 0.3892 |  |

Abbreviations: AMS, antibiotic stewardship; ASP, antibiotic stewardship programme; IPC, infection prevention and control
Data are summarised in Table 3
Item #9 and 10 were excluded from the analysis, as explained in Results.

**Table S3.**
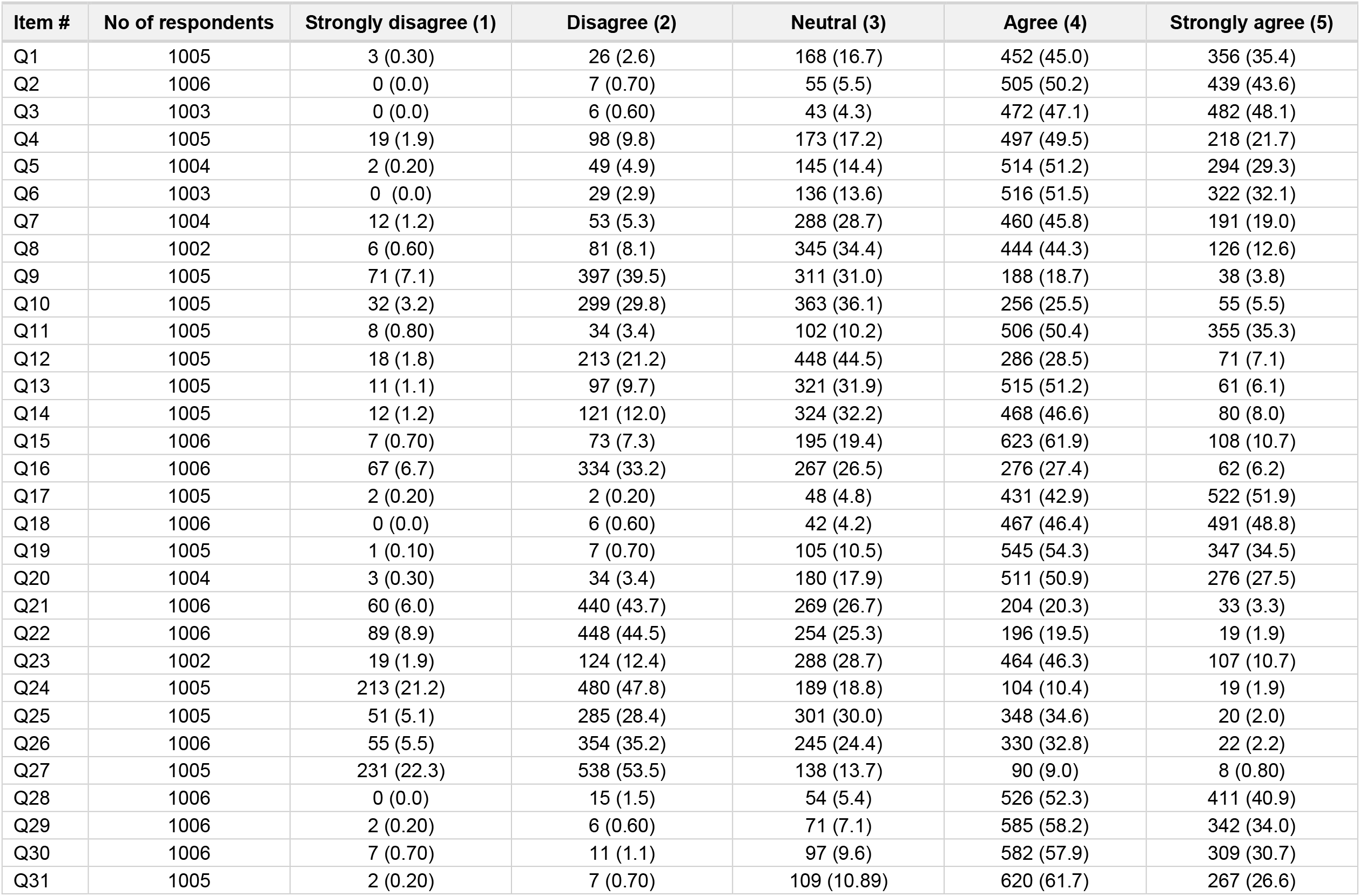
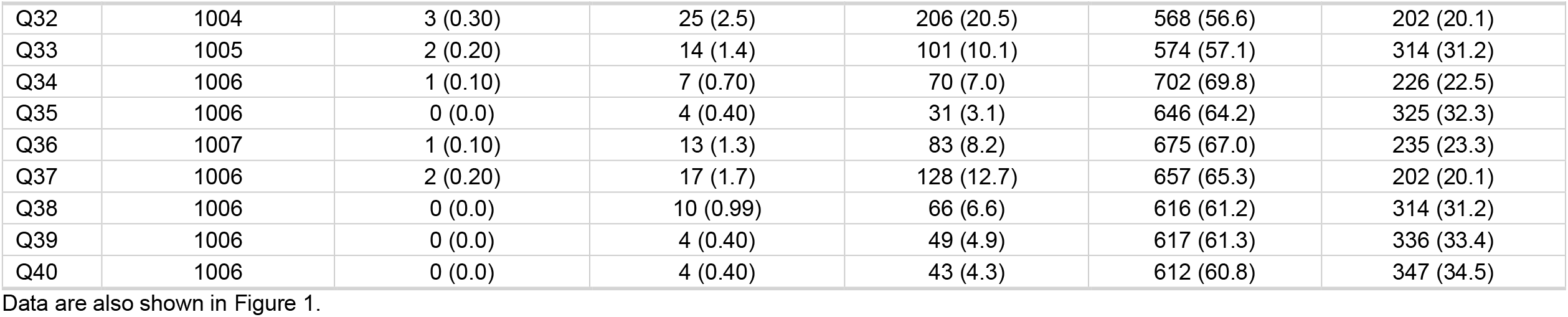
Five-point Likert scale responses for the 40-item questionnaire

**Figure S1.**
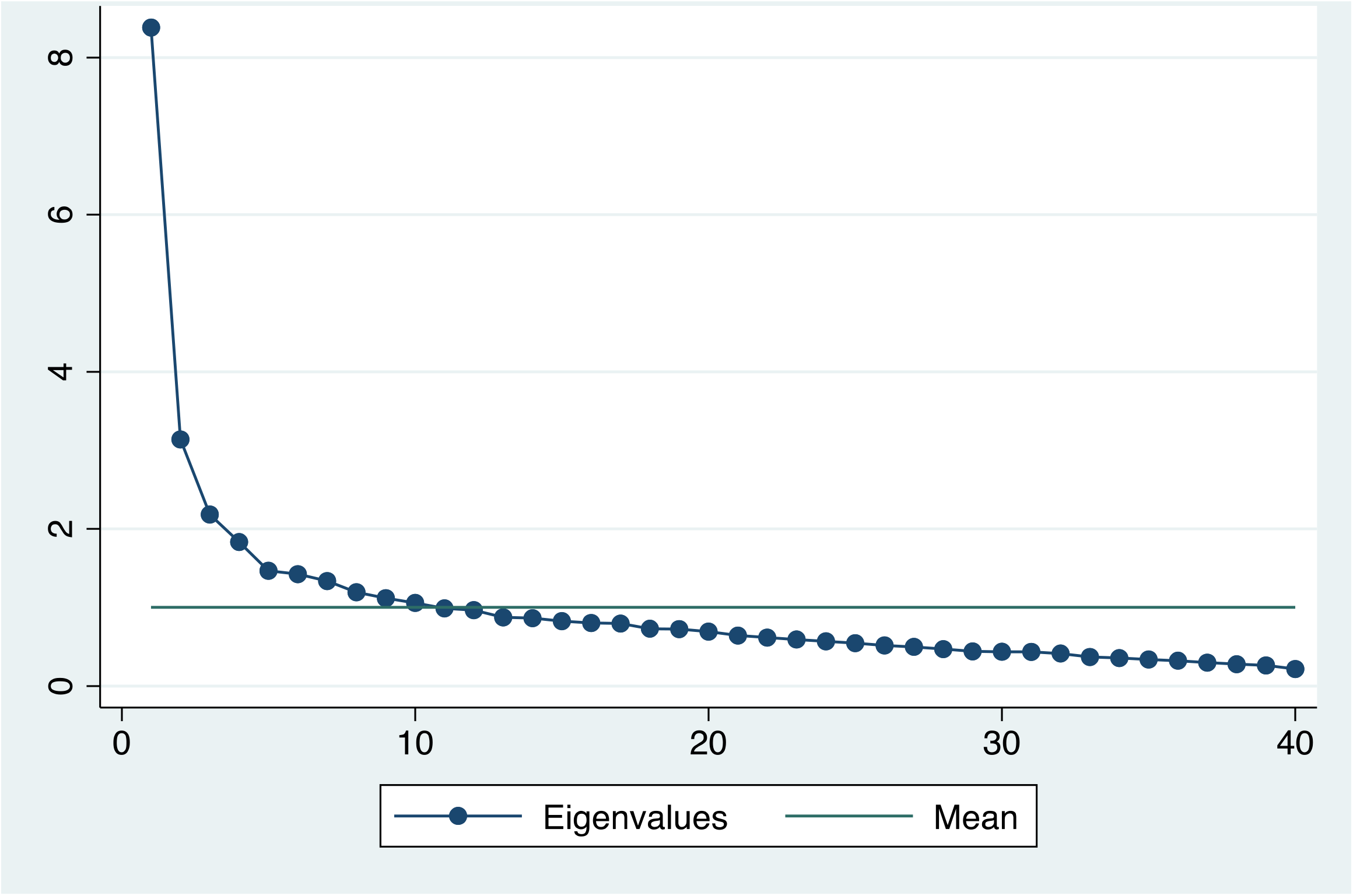
Scree plot showing eigenvalues for the 40 factors

**Figure S2.**
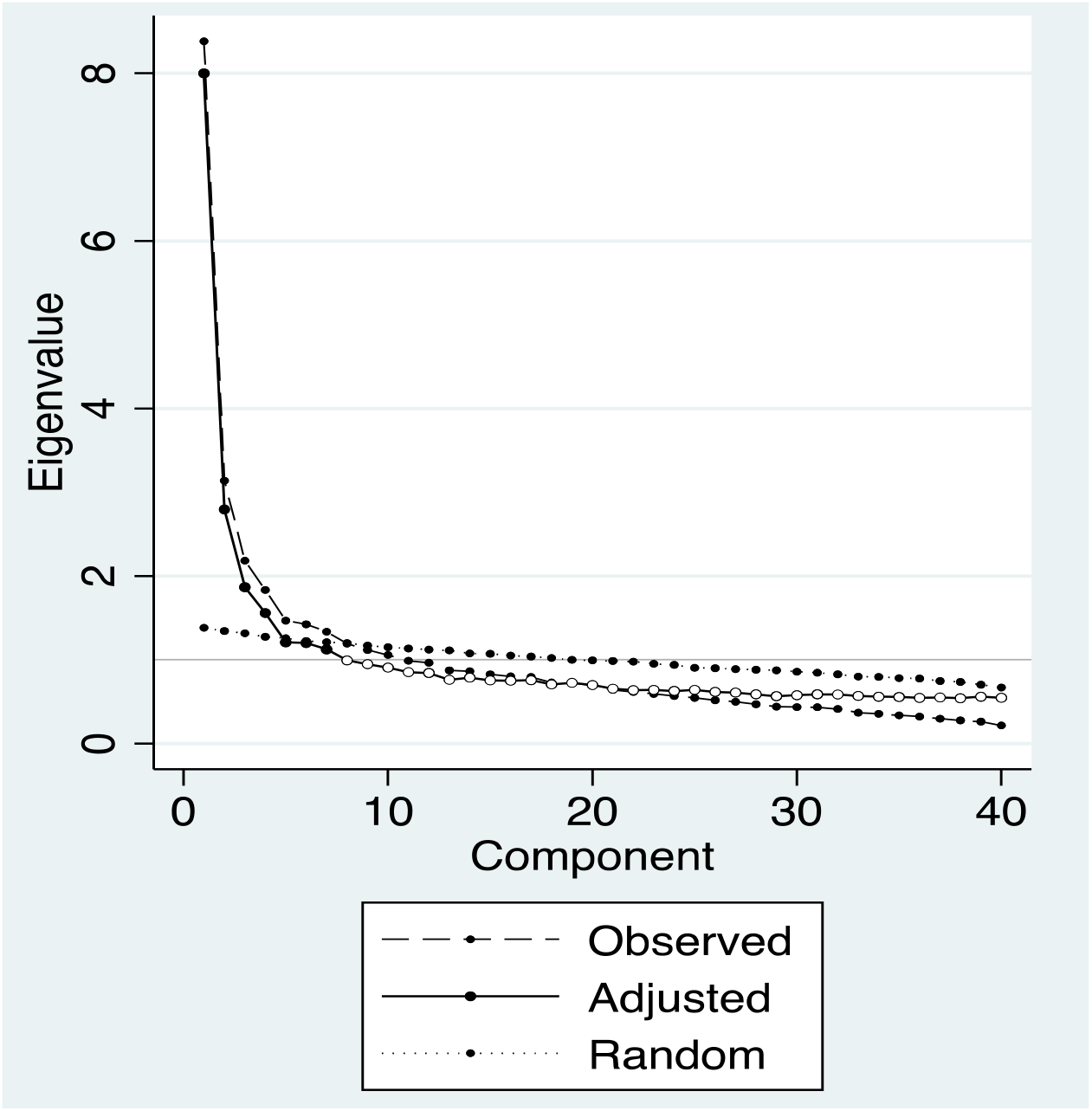
Parallel analysis showing adjusted eigenvalues for the 40 factors. Parallel analysis adjusted the original eigenvalues for sampling error-induced collinearity among the variables to arrive at the adjusted eigenvalues.

